# A Model-Based Meta-Analysis of Pembrolizumab Effects on Patient-Reported Quality of Life: Advancing Patient-Centered Oncology Drug Development

**DOI:** 10.1101/2025.06.14.25329623

**Authors:** Yiqin Zou, Yimeng Sun, Sudhamshu Ravva, Lynne I. Wagner, Jiawei Zhou

**Affiliations:** Division of Pharmacotherapy and Experimental Therapeutics, Eshelman School of Pharmacy, University of North Carolina at Chapel Hill, Chapel Hill, North Carolina, USA; Princeton International School of Mathematics and Science, Princeton, New Jersey, USA; School of Public Health, University of North Carolina at Chapel Hill, Chapel Hill, North Carolina, USA; Lineberger Comprehensive Cancer Center, School of Medicine, University of North Carolina at Chapel Hill, Chapel Hill, North Carolina, USA

## Abstract

Pembrolizumab is an immune checkpoint inhibitor that has been approved for more than 20 different indications and has shown great survival benefits in various types of cancer. However, the reported benefits of pembrolizumab in patients’ quality of life (QoL) have been inconsistent across different studies and different types of cancer. As oncology drug development increasingly emphasizes patient-centered care, patient-reported outcomes (PROs), particularly patient-reported QoL, are recognized as important clinical endpoints. To characterize the effects of pembrolizumab on patient-reported QoL, we conducted a model-based meta-analysis (MBMA) of published clinical trials evaluating pembrolizumab across different types of cancer. The longitudinal EORTC QLQ-C30 GHS/QOL data were extracted in our analysis as QoL scores. A population nonlinear mixed effect (NLME) model was developed to characterize the longitudinal QoL trajectories over time and quantify both treatment toxicity and efficacy. Out of more than 300 screened studies, only 20 reported longitudinal EORTC QLQ-C30 QoL data. Among these, 8 studies reported no between-group differences in QoL outcomes between pembrolizumab and control arms. However, our modeling revealed that pembrolizumab was associated with greater toxicity but improved long-term QoL. Notably, our approach identified treatment effects on QoL that were not detected by traditional statistical analyses in the original publications. In summary, our study demonstrates that MBMA combined with population NLME modeling enables more accurate evaluation of longitudinal PROs data, overcoming the limitations of conventional methods. This approach offers a robust framework for integrating patient-centered outcomes into oncology drug development and supports the broader use of PROs data in regulatory and clinical decision-making.

**Study Highlights:** *What is the current knowledge on the topic?:* Patient-reported outcomes (PROs), particularly patient-reported quality of life (QoL) measures, are essential for patient-centered oncology care. However, PROs data are often underutilized in clinical decision-making due to their subjective nature, high variability, and limited alignment with objective clinical endpoints. Pembrolizumab, a PD-1 immune checkpoint inhibitor, has demonstrated significant survival benefits across multiple cancer types. Yet, many clinical trials have reported no significant improvements in QoL with pembrolizumab treatment.

*What question did this study address?:* This study addressed the questions whether pembrolizumab improves patient-reported QoL and whether population nonlinear mixed-effects (NLME) model can uncover treatment benefits on QoL that may be missed by traditional statistical approaches.

*What this study adds to our knowledge?:* This model-based meta-analysis study shows that NLME modeling approach effectively characterizes treatment effects of pembrolizumab on patient-reported QoL by leveraging longitudinal data. It also highlights the limitations of conventional analyses and the urgent need for standardized PRO instruments and reporting practices.

*How this might change clinical pharmacology and therapeutics?:* This study supports integrating NLME modeling into PROs data analysis to enhance the detection of treatment effects, thereby promoting patient-centered decision-making in oncology drug development and informing future regulatory evaluations.

## Introduction

As oncology drug development continues to evolve, there is a growing emphasis on patient-centered care. Nowadays, treatment success is measured not only by survival outcomes but also by improvements in patient quality of life (QoL).(1) Patient-reported outcomes (PROs), particularly those assessing QoL, are recognized as essential endpoints in cancer clinical trials. (2, 3) These outcomes provide a direct, patient-centered view of treatment benefits, symptom burden, and overall well-being. (4) Despite its clinical importance, PROs are often underutilized in regulatory and clinical decision-making due to the high variability and missingness, subjective reporting, lack of standardization instruments, and inconsistent alignment with clinical survival endpoints. (5–7)

Pembrolizumab, a PD-1 immune checkpoint inhibitor approved for over 20 indications, has shown significant survival benefits across many cancers. (8) Yet, its impact on patient-reported QoL remains unclear. Many trials have reported no significant differences in QoL between pembrolizumab and control groups, despite clear survival advantages. (9–11) This inconsistency may be explained by the high variability in patient-reported data, which limits the power of traditional hypothesis testing. (12) Additionally, most studies only assessed changes from baseline to endpoint, overlooking the importance of longitudinal trajectories of QoL. Prior research suggests that QoL trajectories are more strongly associated with survival than single time-point assessments. (13, 14)

To better understand the effects of pembrolizumab on patient-reported QoL, we conducted a model-based meta-analysis (MBMA) of published clinical trials across multiple cancer types. We applied population nonlinear mixed effect (NLME) modeling to analyze longitudinal patient-reported QoL data for pembrolizumab, accounting for both between-study and between-arm variability. This approach enables a more precise assessment of pembrolizumab’s benefits on QoL. Our analysis also highlights the potential of model-based methods to improve the evaluation of PROs in oncology drug development. By providing a scalable and quantitative framework for analyzing PROs data, this study supports the broader adoption of patient-centered endpoints in drug evaluation, regulatory review, and clinical decision-making.

## Methods

### Literature Search

We conducted a PubMed search (http://www.ncbi.nlm.nih.gov/pubmed/) using the query: *((Pembrolizumab) AND (melanoma)) AND (patient-reported OR quality of life OR PRO-CTCAE)*, with the filter “Clinical Trial” to include only clinical studies reporting patient-reported outcomes (PRO) for pembrolizumab in melanoma. The term “melanoma” was then replaced with other cancer types, including non-small-cell lung cancer (NSCLC), triple-negative breast cancer (TNBC), head and neck squamous cell carcinoma (HNSCC), urothelial cancer, and endometrial/colorectal cancer (CRC), to identify studies across multiple indications. Studies without longitudinal PRO data were excluded. We further restricted the final database to studies reporting longitudinal EORTC QLQ-C30 Global Health Status/Quality of Life (GHS/QoL) scores, as this is the most widely used PRO instrument in oncology clinical trials. (15) The GHS/QOL scores (referred to as QoL scores in this study) are derived from two items in the EORTC QLQ-C30 questionnaire:

1. How would you rate your overall health during the past week?
2. How would you rate your overall quality of life during the past week?

Each item is rated on a 7-point scale ranging from 1 (very poor) to 7 (excellent). The average of the two scores is then linearly transformed according to EORTC scoring guidelines. (16) The resulting longitudinal QoL scores range from 0 to 100, with higher scores indicating better overall health status and quality of life.

### Data Extraction

Longitudinal QoL scores from either pembrolizumab or control arm were extracted from published figures or tables using WebPlotDigitizer (https://automeris.io/). All included data were required to have at least three QoL measurements per arm; studies reporting only baseline and endpoint QoL scores were ineligible in our analyses. Variability measures associated with the QoL scores were digitized and recorded when available. Additional trial-level information including study design, sample size, patient age, gender, tumor type, disease stage, and Eastern Cooperative Oncology Group (ECOG) performance status was also collected to construct MBMA database. All data were collected as aggregate-level summaries by study arm.

### Longitudinal QoL Trajectory Structural Model Development

To characterize the longitudinal trajectories of QoL scores, we developed a semi-mechanistic population model using data from the MBMA database. In this model, treatment-related side effects were captured as a decline in QoL scores using an asymptotic exponential function, characterized by two parameters: *EMAX* (maximum toxicity) and *Kp* (rate constant for toxicity onset). The treatment-related improvement in overall well-being and the decline in QoL due to disease progression were jointly modeled using a linear function with a slope parameter (*SLP*). The QoL trajectory structural model is presented in **Equation 1**.

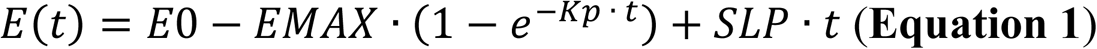

Where *E(t)* is the QoL scores change over time, *E0* is the QoL score at baseline, *SLP* is the QoL improvement rate, *EMAX* is the maximal toxicity, and *Kp* is the toxicity offsite rate (1/week), *t* is the time in week.

### Random Effects Model Development

To ensure that model-predicted QoL scores stayed within the 0 to 100 range, a logit transformation was applied to the observed scores as the model input. The transformed value is defined in **Equation 2**:

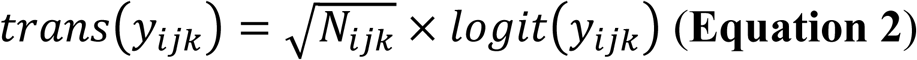

Where *y*_*ijk*_ is the observed QoL scores in study *i,* at the time point *j*, and treatment arm *k*. The term *trans*(*y*_*ijk*_) represents the logit-transformed *y*_*ijk*_ used as the dependent variable (DV) in the model. *N*_*ijk*_ is the number of individuals corresponding to that observation.

A constant residual error weighted by sample size *N*_*ijk*_ was used to describe residual variability in **Equation 3**.

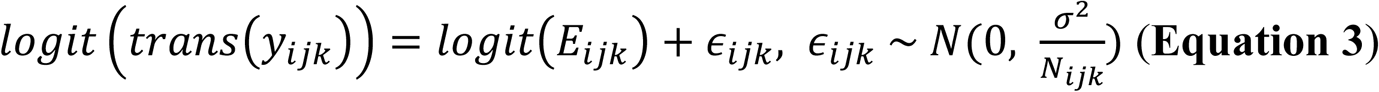

Where *E*_*ijk*_ is the model-predicted transformed QoL, and *∈*_*ijk*_ is the constant residual error following a normal distribution with mean 0 and variance *σ*^2^ scaled by sample size *N*_*ijk*_.

To capture heterogeneity across studies and treatment arms, between-study variability (BSV) and between-treatment arm variability (BTAV) were incorporated into model parameters. For parameters bounded between 0 and 100, such as baseline QoL score *E0*, a logit transformation was applied to map them to the continuous parameter space when estimating random effects, as shown in **Equation 4**:

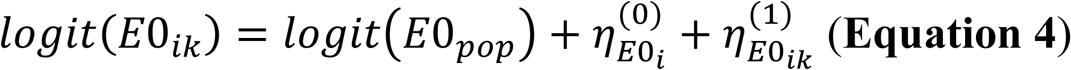

Where *E*0_*ik*_ is the baseline QoL scores for study *i* and treatment arm *k*. The *E*0_*pop*_ is the population parameter for *E0*. The term 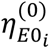 and 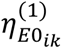 represents the BSV and BTAV following a normal distribution with mean 0 and standard deviation *Ω*_*BSVE*0_ and *Ω*_*BTAE*0_, respectively.

Parameter estimation was performed using a nonlinear mixed-effects (NLME) modeling approach implemented via the Stochastic Approximation Expectation-Maximization (SAEM) algorithm with linear method.

### Covariate Model Development

The effects of pembrolizumab or control arm on longitudinal QoL trajectories were modeled as covariates using an exponential function (**Equation 5**):

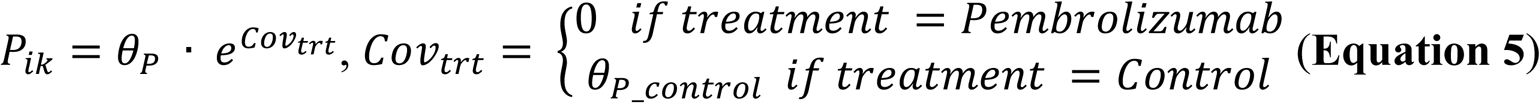

Where *P_ik_* is the individual value for parameter *P* in study *i* arm *k*, and *θ_P_* is the typical value for parameter *P*, *θ_P_control_* is the estimable parameter for the effect of control arm relative to pembrolizumab on parameter *P*.

In addition to treatment effects, other baseline covariates were evaluated based on exploratory analysis and clinical relevance. These covariates included age, proportion of male patients, proportion with stage IV disease, proportion with ECOG performance status of 0, and tumor type. Covariates were explored for their potential influence on key model parameters *E0*, *Emax*, and *SLP*.

A full fixed-effects modeling approach was used, in which all relevant covariates were added to the base model simultaneously. Covariate correlations were assessed, and when strong correlations were present, the more clinically relevant or statistically significant covariate was retained. Categorical covariate effects were modeled as shown in **Equation 5**. Continuous covariates were modeled using an exponential function (**Equation 6**). For example, the effect of Age on parameter *P* was described as:

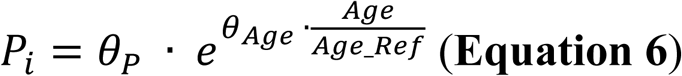

Where *θ_Age_* is the estimable parameter for the effect of age, and *Age_Ref* is the reference value for age.

Missing values for covariates were imputed with the population median (for continuous covariates) or mode (for categorical covariates) during the covariate analyses.

### Assessment of Model Performance

Model performance was evaluated based on changes in the objective function value (OFV), diagnostic plots, visual predictive check (VPC), and assessments of parameters precision and shrinkage. Diagnostic plots included observations versus population predictions or individual predictions, individual weighted residuals versus time or predictions, and individual predictions over time overlaid with observations. (17) VPCs were conducted using 1,000 simulations to compare observed data with model-predicted intervals, assessing the model’s ability to capture both central trends and variability in the data. (18) Parameter precision was evaluated using relative standard error (RSE), with values below 30% indicating reliable and stable estimates. Shrinkage, calculated for parameters with interindividual variability (IIV), reflects how much individual estimates are pulled toward the population mean. Shrinkage below 30% suggests adequate capture of BSV and BTAV, while higher shrinkage may indicate unreliable individual predictions even when overall diagnostics appear acceptable. (19, 20)

### Model Selection

Several structural models were examined during model development, including linear and exponential forms of QoL improvement, with or without treatment-related toxicity. Different configurations of BSV and BTAV were tested across model parameters. Each candidate model was assessed using diagnostic plots, VPC, and parameter estimates. The final model was selected based on better goodness-of-fit, improved VPC performance, acceptable parameter precision and shrinkage, and a lower OFV.

### Model Validation

To assess model stability and parameter uncertainty, nonparametric bootstrap validation was conducted with 500 replicates. In each replicate, a new dataset was generated by random sampling with replacement from the original dataset at the study arm level, preserving the longitudinal structure within each arm. The model was re-estimated for each bootstrap replicate using the same structural and statistical model. The distribution of parameter estimates across replicates was used to construct parameter 10^th^/90^th^ percentiles, evaluate estimation robustness, and assess the precision of fixed-effect and random-effect parameters.

### Mode Simulation

The longitudinal impact of pembrolizumab on QoL trajectories was evaluated using model-based simulations and compared with control arm. A total of 500 simulations were conducted, incorporating BSV and BTAV to capture individual heterogeneity.

### Statistical Analysis

The effects of treatment type (pembrolizumab versus control) on QoL trajectory parameters will be compared using Wilcoxon tests. A two-sided p-value < 0.05 will be considered statistically significant.

### Software and Code Availability

MBMA model was developed using Monolix 2024R1. The model simulations were performed using Simulx 2024R1. Both Monolix and Simulx can be downloaded at https://lixoft.com/products/. Plots and survival analyses were performed using R 4.4.1 and RStudio Version 2022.07.1+554. The figures were reorganized in GraphPad Prism (Version 10.4.1). The model codes are provided in the **Supplementary Codes**.

### Data Availability

The data was accessed through published literature. The organized MBMA database is available from the corresponding author upon reasonable request.

## Results

### MBMA Database

Initial literature screening was completed by March 16, 2025, identifying 302 studies across 7 tumor types. Studies were included if they reported longitudinal QoL scores, resulting in 20 studies selected for final inclusion in the MBMA database. The inclusion and exclusion processes are illustrated in **Figure S1**. Study information, along with the number of available QoL data points and sample sizes, is summarized in **Table 1**. Of the studies comparing pembrolizumab to control, 8 reported improved QoL with pembrolizumab, while 8 found no significant difference between groups. In total, 410 QoL data points were collected, with 228 from pembrolizumab arms and 182 from control arms. The aggregated longitudinal QoL trajectories are shown in **Figure 1A**. To assess potential bias, we compared the QoL effect size between pembrolizumab and control arm against data variability using a funnel plot. (**Figure 1B**) Most data points fell within the 95% confidence interval, indicating no apparent publication or selection bias in the dataset.

**Figure 1.**
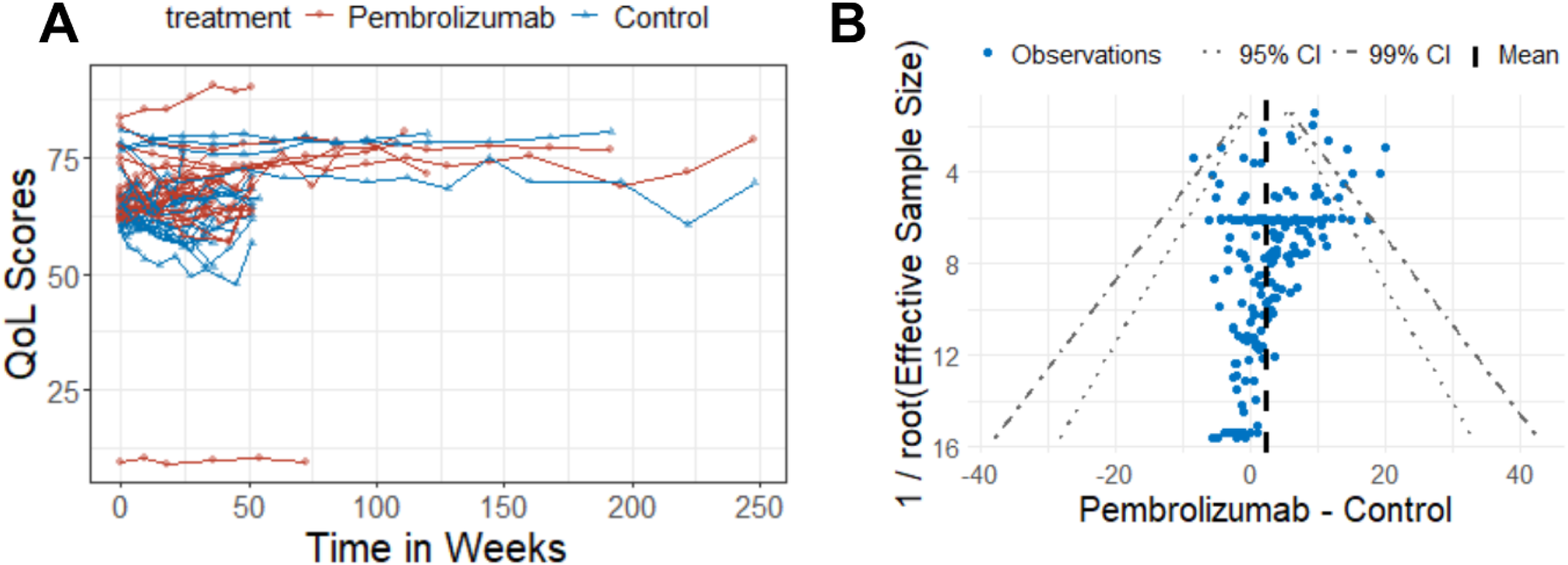
Data visualization and assessment of data bias using a funnel plot. (A) Longitudinal trends in EORTC QLQ-C30 GHS/QOL scores for pembrolizumab and control arms. Each circle represents an arm-level aggregate data point; each line connects repeated measures within the same arm over time. (B) Funnel plot comparing the difference in QoL scores between pembrolizumab and control arms against the inverse square root of effective sample size. The solid black dashed line indicates the mean effect size, with 95% and 99% confidence intervals shown using different dashed line styles. QoL, quality of life; TRT, treatment; CI, confidence interval; EORTC QLQ-C30, European Organisation for Research and Treatment of Cancer Quality of Life Questionnaire-Core 30; GHS/QOL, Global Health Status / Quality of Life.

**Table 1.**
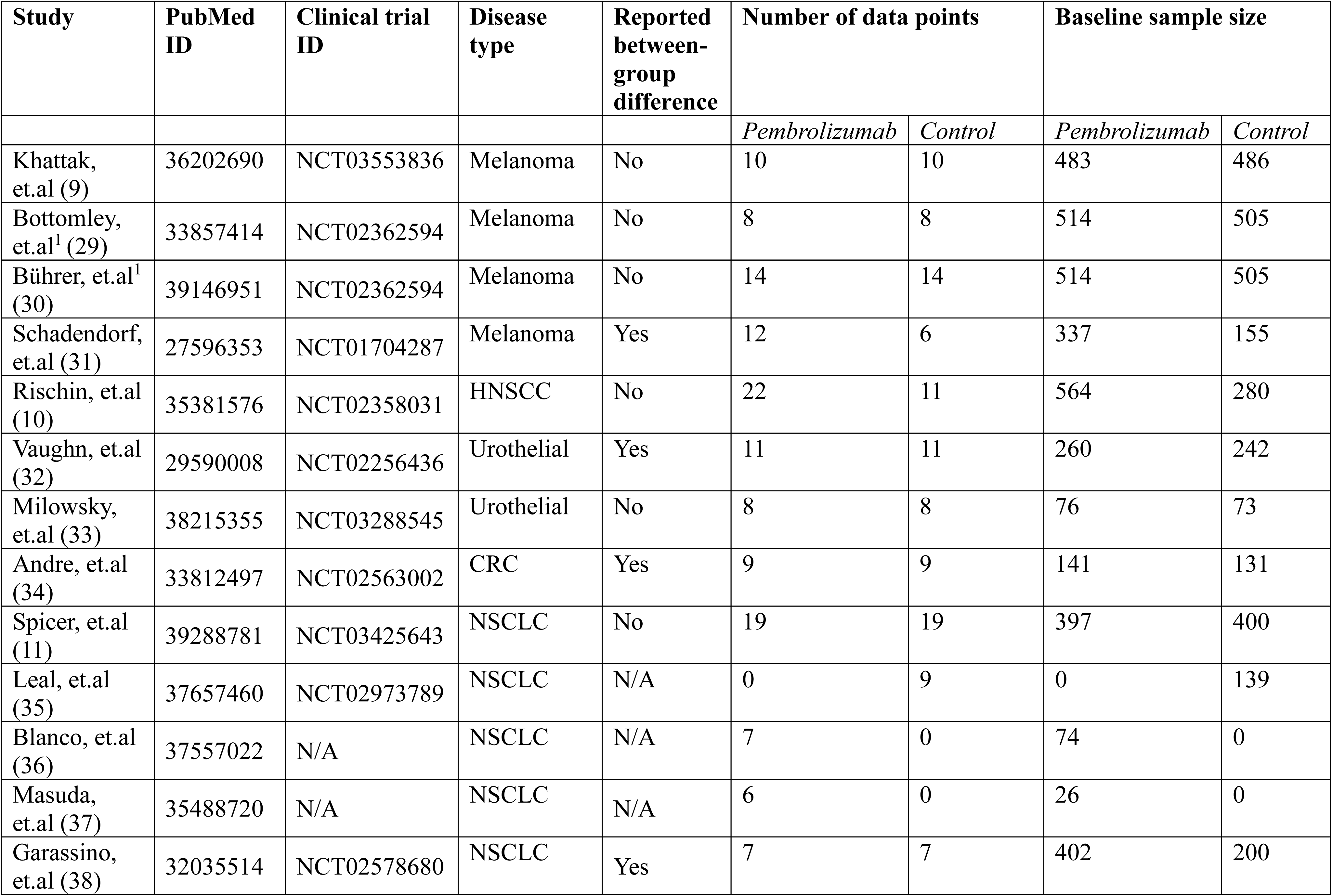
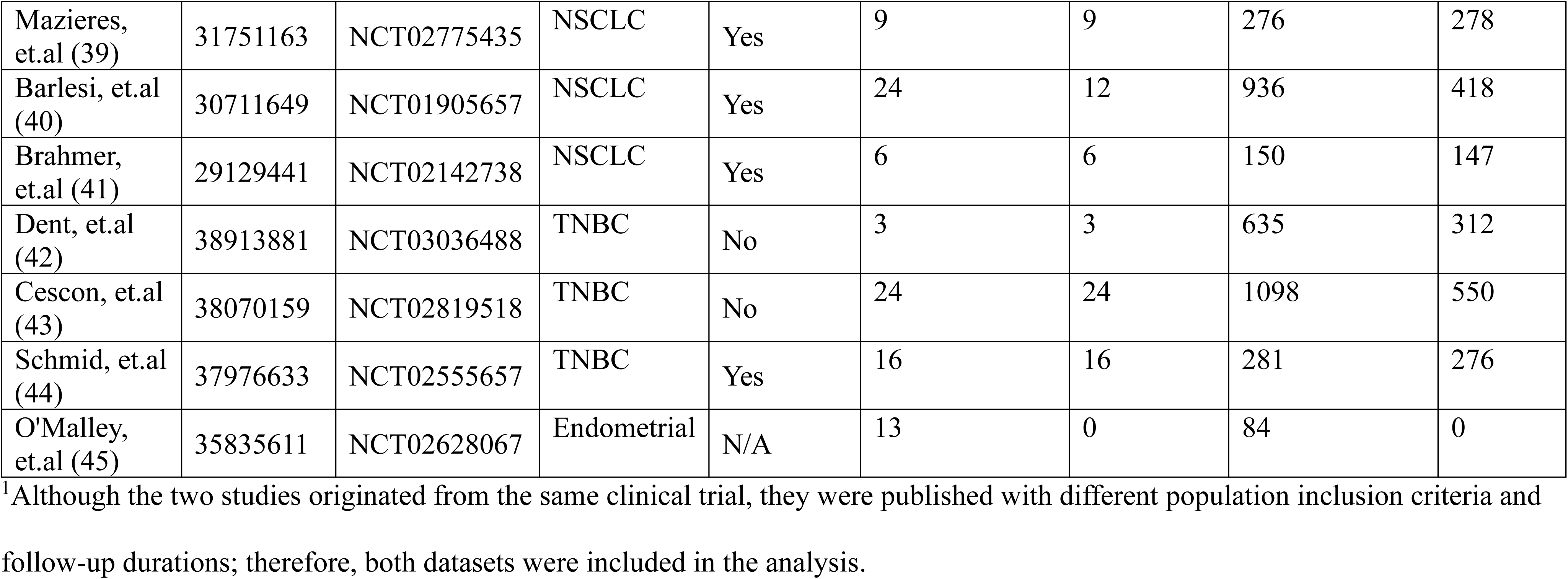
Summary of included studies and analyzed data.

### MBMA Model Analysis

The model describing longitudinal QoL trajectories was developed using the MBMA database and demonstrated a good fit to the data. Final model diagnostic plots showed no apparent bias in either the pembrolizumab or control arms. (**Figure 2A**) The VPC plot indicated that the model’s prediction intervals adequately captured the observed data, supporting strong model performance. (**Figure 2B**) **Figure 3** presents observed QoL scores over time alongside model-predicted trajectories for each individual study. Despite the high variability in the data, the model successfully captured the overall trends and variability in QoL trajectories.

**Figure 2.**
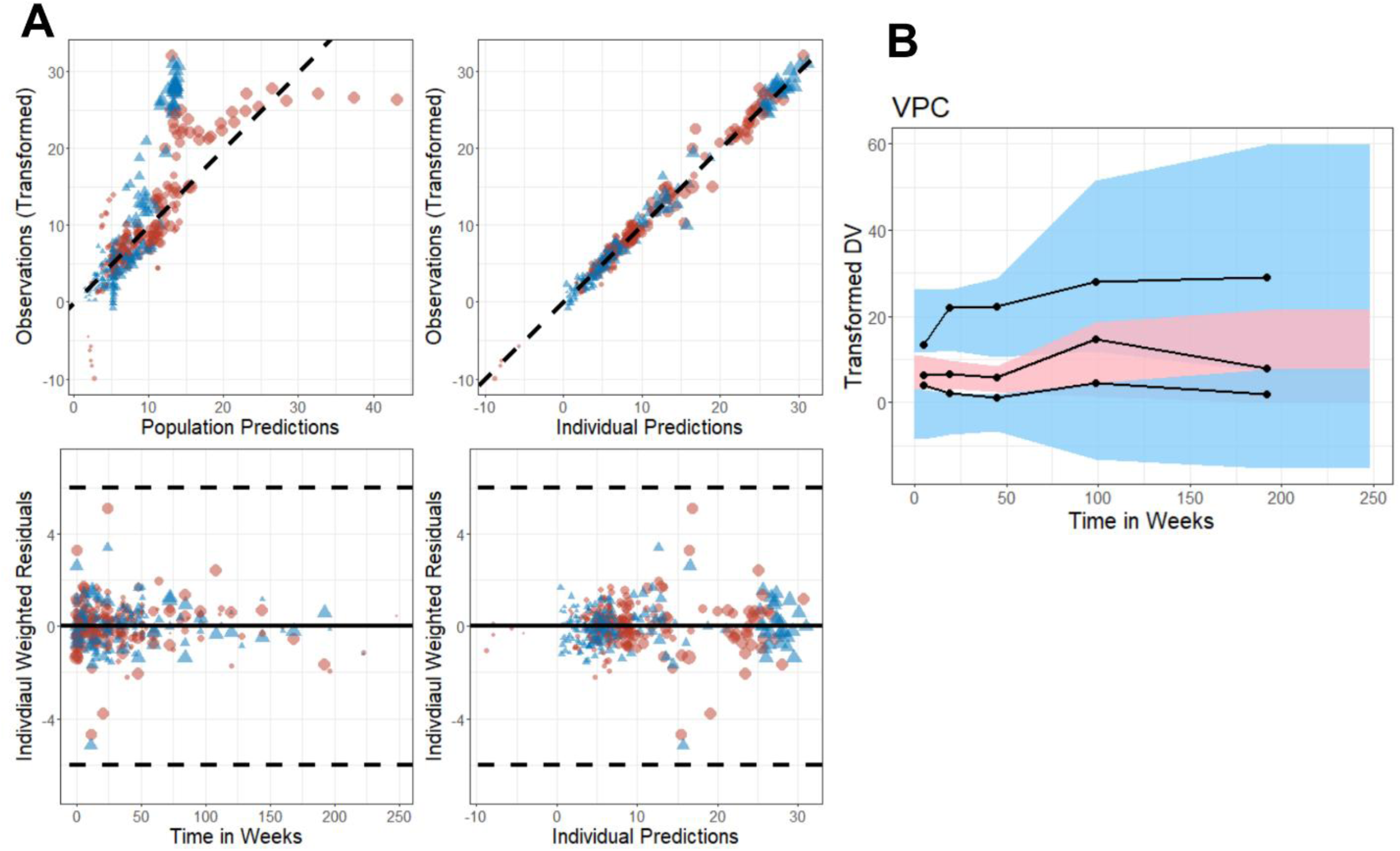
Final model diagnostics plots and VPC. (A) Diagnostic plots include: model-predicted population values vs. transformed observations (top left), and individual predictions vs. transformed observations (top right). The black dashed line represents the identity line (y = x). Residual plots show individual weighted residuals vs. time (bottom left) and vs. individual predictions (bottom right), with the black solid line at y = 0 and dashed lines at y = ±6. Red circles represent pembrolizumab data and blue triangles represent control arm data; symbol size reflects arm sample size. (B) VPC of transformed QoL scores over time. Observed data are shown as black solid lines (median and 10th/90th percentiles), and model-predicted data from 1000 simulations are shown as shaded areas: red (90% PI of the median) and blue (90% PI of the 10th and 90th percentiles). VPC, visual predictive check; PI, prediction interval; QoL, quality of life.

**Figure 3.**
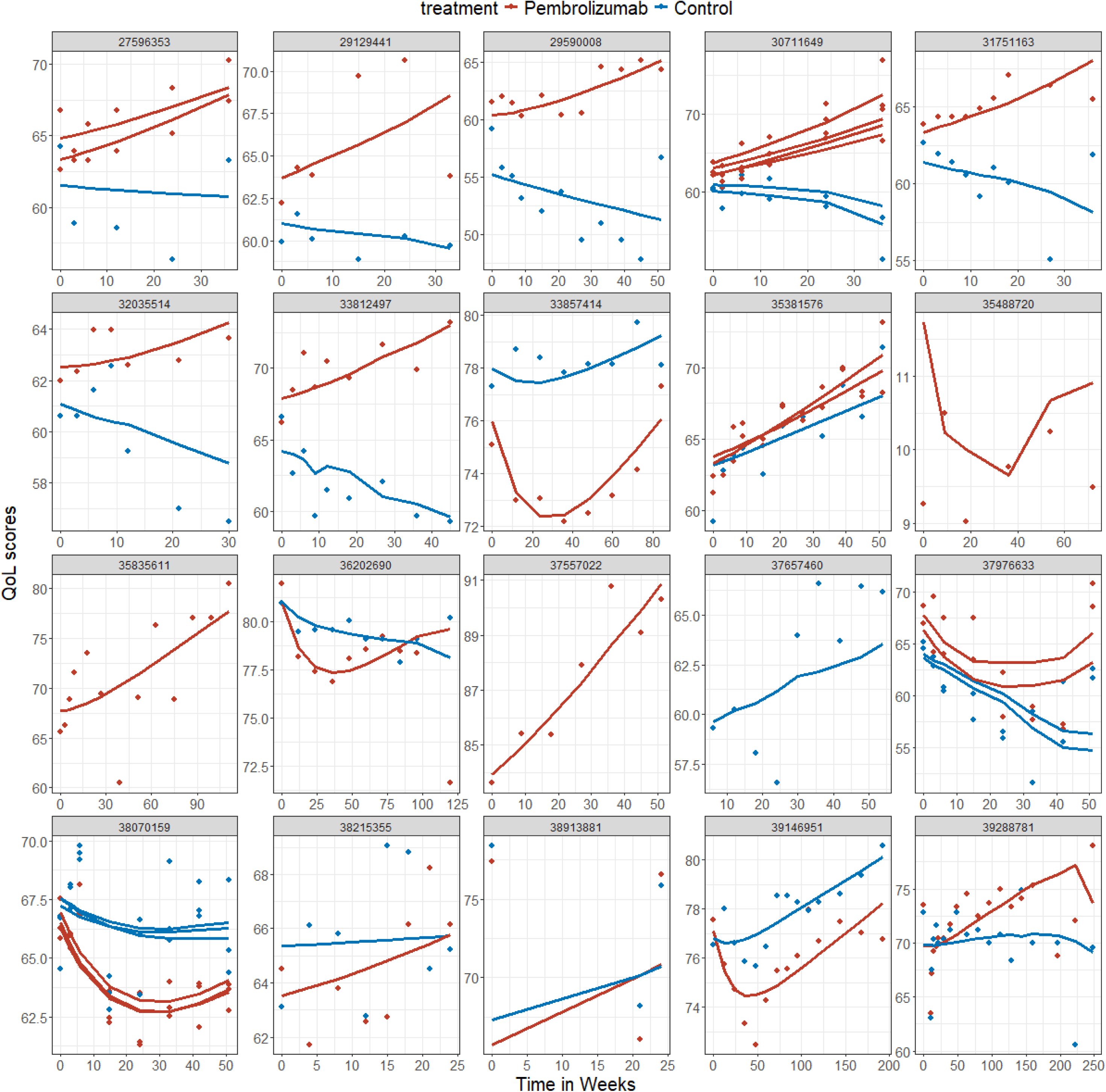
Goodness-of-fit by individual study. Each subpanel represents a study (labeled by PubMed ID). Dots indicate observed QoL scores over time, while solid lines represent model-predicted individual QoL trajectories. Data are stratified by treatment arm. QoL, quality of life.

In the final MBMA model, treatment effect was identified as a significant covariate on maximal toxicity (*Emax*) and QoL improvement rate (*SLP*). No additional significant covariates were identified during model development. Parameter estimates from the final model, along with bootstrap results, are presented in **Table S1**. All parameter estimates had reasonable RSE, indicating precise and robust estimation. Shrinkage values were all below 30%, suggesting good characterization of between-study and between-arm variability.

### MBMA reveals pembrolizumab longitudinal QoL benefits

Using our developed MBMA model, we simulated longitudinal QoL trajectories for patients receiving pembrolizumab versus control. **(Figure 4A**) Patients treated with pembrolizumab experienced greater initial toxicity but demonstrated sustained long-term improvements in QoL scores.

**Figure 4.**
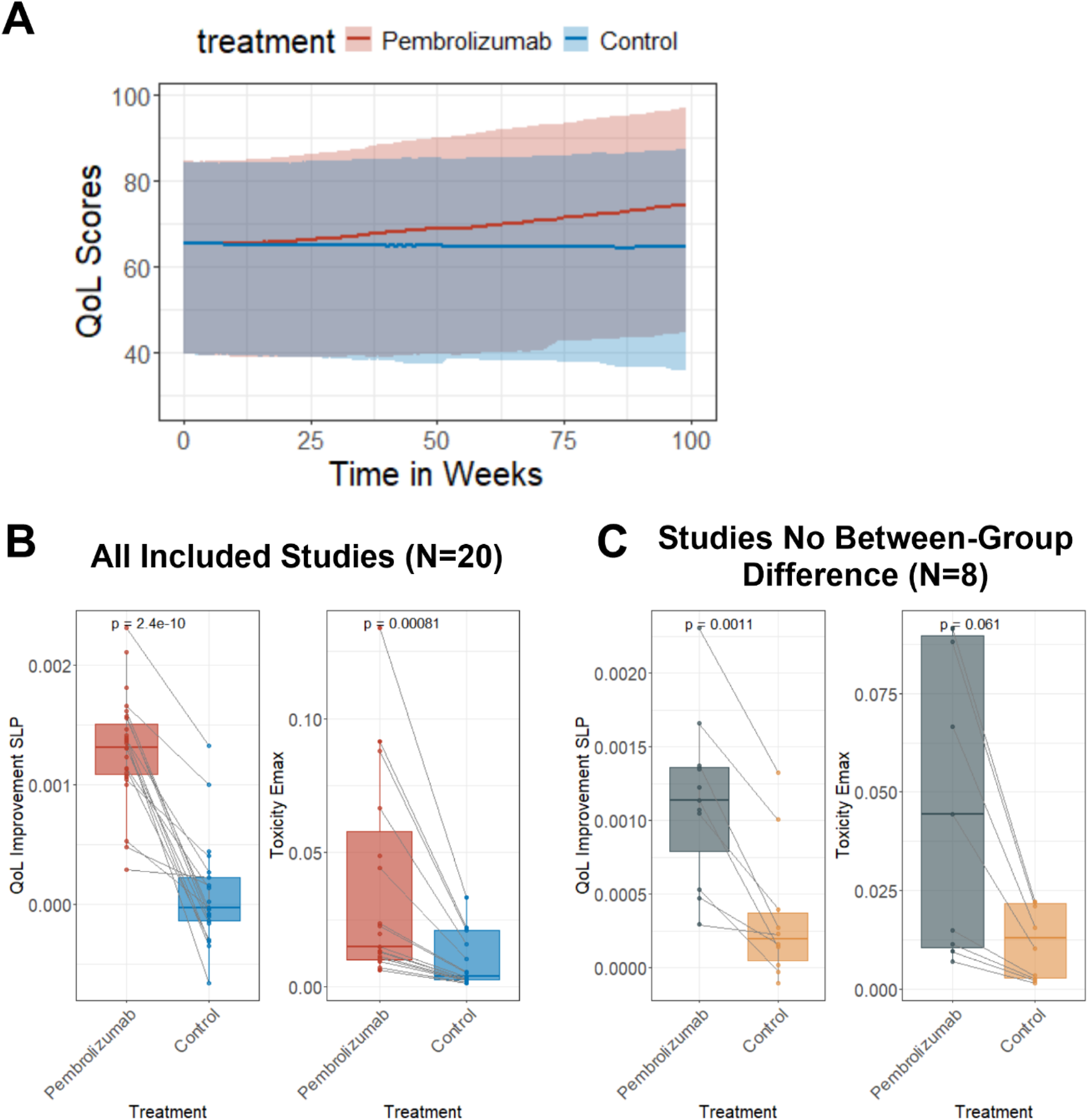
Longitudinal quality of life (QoL) trajectory comparisons between pembrolizumab and control arms. Model estimated QoL trajectory parameters, including QoL improvement slope (*SLP*) and maximal toxicity (*Emax*), were compared for all included studies (A) and for studies that reported no between-group differences in the original publication (B). (C) Longitudinal model simulated QoL scores trajectories in pembrolizumab or control arms.

To compare longitudinal QoL trajectories between pembrolizumab and control arms, we selected two key MBMA model parameters: the QoL improvement rate (*SLP*) and maximal toxicity (*Emax*). Pembrolizumab was associated with a significantly higher QoL improvement rate (*SLP*, p < 0.0001) and significantly greater initial toxicity (*Emax*, p = 0.005), compared to control arm (**Figure 4B**). These findings are consistent with prior clinical observations that immune checkpoint inhibitors, such as pembrolizumab, may induce early toxicities (e.g., fatigue, diarrhea, rash) that negatively impact QoL, while offering longer-term clinical benefits such as prolonged survival and delayed disease progression. (21, 22)

We further conducted a subgroup analysis focusing on studies that originally reported no between-group differences in QoL. (**Table 1**) Even within this subset, the model-estimated QoL improvement rate (*SLP*) remained significantly higher in the pembrolizumab arm compared to control (p < 0.05), and a borderline significantly higher toxicity (*Emax*) was observed (p = 0.052) in pembrolizumab. (**Figure 4C**) These results highlight the strength of the NLME modeling approach in accounting for variability in PRO data and capturing longitudinal treatment effects that may be missed by conventional statistical comparisons. Incorporating the full trajectory of longitudinal PRO data, rather than relying solely on baseline-to-endpoint comparisons, may offer a more sensitive and informative assessment of treatment impact on patient QoL.

## Discussion

As oncology drug development continues to shift from a disease-centered to a patient-centered paradigm, it is essential to ensure that therapies not only prolong patient survival but also improve patients’ QoL. (13, 23) In this study, we conducted an MBMA study to evaluate patient-reported QoL outcomes associated with pembrolizumab across multiple cancer types. To address the substantial variability commonly seen in patient-reported QoL data, which often limits the effectiveness of traditional statistical methods, we applied NLME modeling approach. This method allowed us to incorporate the longitudinal trajectory of QoL data and was found to more accurately capture treatment benefits compared to the statistical approaches used in the original publications. Our analysis showed that although pembrolizumab was associated with greater initial toxicity compared to control, it provided meaningful long-term improvements in QoL across different cancer populations. These findings offer a strong quantitative foundation to guide more patient-focused drug development and inform future regulatory decisions.

MBMA offers a robust quantitative framework for integrating data across multiple clinical trials while accounting for between-study heterogeneity. (24) In our MBMA study, an initial literature search identified over 300 publications reporting pembrolizumab efficacy data. However, only 6.6% (N = 20) of these studies included valid longitudinal patient-reported QoL data. Most publications were excluded due to the absence of PROs or the use of non-standardized instruments to assess QoL. Although several guidelines have emphasized the importance of PROs (25, 26) and the FDA has recommended their inclusion in early-phase oncology trials to support dose optimization and patient-centered treatment (27), our findings reveal a persistent lack of standardized PRO instruments and data reporting. This gap highlights the urgent need to improve and standardize PRO collection and reporting practices in future clinical trials to better inform decisions made by patients, clinicians, and regulatory stakeholders.

The PRO data from Phase III clinical trials are rarely incorporated into clinical decision-making, often due to the lack of significant between-group differences—despite clear survival benefits. A review of drug approvals between 2011 and 2017 found that only 10 out of 71 (14%) showed PRO benefits at the time of approval. (28) This has limited the use of PRO data in regulatory and clinical decision-making. In this study, we used a population NLME modeling approach to analyze longitudinal QoL trajectories and characterize treatment effects on PRO endpoints. Compared to traditional hypothesis tests, the NLME approach more effectively detected treatment-related differences by incorporating the full trajectory of longitudinal data and quantifying interindividual variability. Our findings demonstrate the value of applying population modeling to PRO analysis and highlight its potential to enhance the integration of patient-centered outcomes into drug development and decision-making.

Our study has limitations. First, the dataset lacked sufficient power to compare pembrolizumab’s QoL benefits across different tumor types. Future research is needed to determine whether pembrolizumab’s toxicity profile and QoL effects differ by cancer type. Another limitation is that our MBMA model was developed on arm-level aggregate data, whereas the statistical analysis on QoL data in the original clinical studies used individual-participant level data. This creates a gap that limits the direct comparison between pharmacometrics and traditional statistical methods. Further analyses using individual participant-level data are needed to validate the utility of population modeling in PRO data analysis.

## Conclusion

In conclusion, we conducted an MBMA to evaluate the impact of pembrolizumab on patient-reported QoL across multiple cancer types. By applying NLME modeling approach, we were able to characterize the longitudinal QoL trajectories and quantify treatment effects that might be confounded by the data variability when using traditional statistical hypothesis tests. Our findings support the use of advanced modeling techniques to improve the interpretation of PRO data and highlight the importance of incorporating longitudinal QoL measures into clinical trial evaluation. Ultimately, this work contributes to a growing body of evidence promoting more patient-centered approaches in oncology drug development and regulatory decision-making.

## Data Availability

All data produced in the present study are available upon reasonable request to the authors

## Acknowledgements

We acknowledge the longitudinal model-based meta-analysis tutorial provided by Lixoft (https://monolixsuite.slp-software.com/tutorials/2024R1/case-study-longitudinal-model-based-meta-analysis-). This work is funded by the University of North Carolina at Chapel Hill.

## Author Contributions

J.Z and L.I.W designed the study and wrote the manuscript. Y.Q and Y.S. collected the data. S.R performed analyses.

## Notes

**Conflicts of Interests:** The authors report no conflicts of interest.

### Competing Interest Statement

The authors have declared no competing interest.

