## Supplementary Materials for "A Model-Based Meta-Analysis of Pembrolizumab Effects on Patient-Reported Quality of Life: Advancing Patient-Centered Oncology Drug Development"

**Table S1. Final model parameter estimates.**

| **Parameter** | **Value** | **RSE (%)** | **Bootstrap median** | **Bootstrap $10^{th}/90^{th}$ Percentile** | **SHR (%)** |
|---|---|---|---|---|---|
| **Population parameters** | | | | | |
| Baseline QoL score (*E0*) | 0.656 | 5.16 | 0.656 | 0.641 – 0.67 | |
| Maximal toxicity reducing QoL (*Emax*) | 0.0268 | 36.2 | 0.023 | 0.011 – 0.04 | |
| Toxicity offset rate (*Kp*, 1/week) | 0.0705 | 30.4 | 0.069 | 0.036 – 0.526 | |
| QoL improvement rate (*SLP*, 1/week) | $9.98 \times 10^{-4}$ | 24.1 | 0.94 | $5.44 \times 10^{-4}$ – $1.498 \times 10^{-3}$ | |
| Effect of control arm on *Emax,* pembrolizumab as reference ($\theta_{Emax\_PBO}$) | -0.758 | 32.6 | -0.642 | -1.399 – -0.006 | |
| Effect of control arm on *SLP,* pembrolizumab as reference ($\theta_{SLP\_PBO}$) | -1.4 | 32.2 | -1.371 | -2.076 – -0.768 | |
| Between-study variability on *Emax* (*BSVEmax*) | 0 Fixed | | | | |
| Between-study variability on *E0* (*BSVE0*) | 0 Fixed | | | | |
| Between-arm variability on *E0* (*BTAE0*) | 0 Fixed | | | | |
| **Inter-individual variability (standard deviation)** | | | | | |
| Standard deviation of *BSVEmax* ($\Omega_{BSVEmax}$) | 0.783 | 31.7 | 0.903 | 0.476 – 1.267 | 17.3 |
| Standard deviation of *BSVE0* ($\Omega_{BSVE0}$) | 0.667 | 16.1 | 0.665 | 0.314 – 1.044 | 29.9 |
| Standard deviation of *BTAE0* ($\Omega_{BTAE0}$) | 0.867 | 18.4 | 0.852 | 0.565 – 1.191 | 20 |
| Standard deviation of *SLP* ($\Omega_{SLP}$) | 0.871 | 16.1 | 0.834 | 0.446 – 1.232 | 25.8 |
| **Residual unexplained variability** | | | | | |
| Additive residual error (***a***) | 1.1 | 3.95 | 1.068 | 0.891 – 1.276 | |

RSE, relative standard error; CI, confidence interval; SHR, shrinkage.

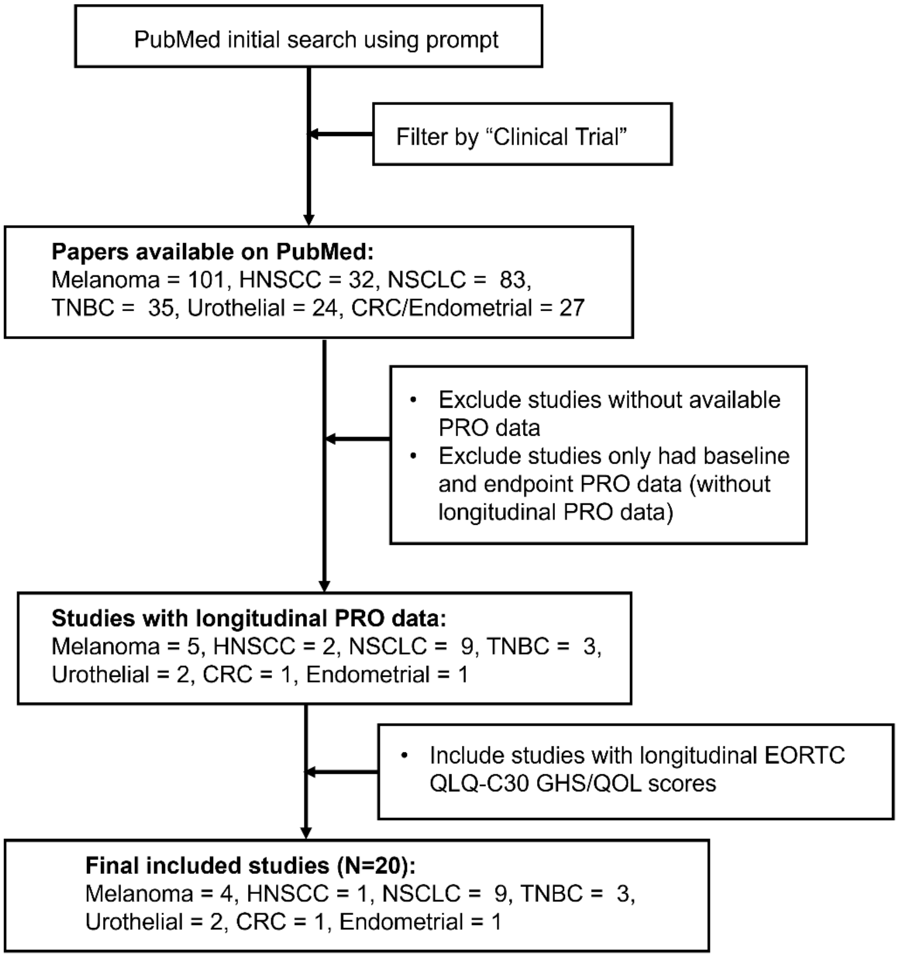


**Figure S1. Workflow diagram of studies included in the analysis.** HNSCC, head and neck squamous cell carcinoma; NSCLC, non-small-cell lung cancer; TNBC, triple-negative breast cancer; CRC, colorectal cancer; PRO, patient-reported outcomes. GHS/QOL, Global Health Status / Quality of Life.

### Supplementary Codes

#### A. MBMA model Monolix codes.

```
DESCRIPTION:
MBMA for QOL.

[LONGITUDINAL]
input = {E0FE, EmaxFE, Kp, SLP,NOC, etaBSVEmax, etaBSVE0, etaBTAVE0 }
NOC    = {use=regressor}

EQUATION:
; Baseline QoL
; transform the E0 fixed effect (E0FE) to tE0 (normally distribute)
tE0 = logit(E0FE)

; adding the random effects (RE) due to between-study variability and
; between arm variability, on the transformed parameters
tE0RE = tE0 + etaBSVE0 + etaBTAVE0/sqrt(NOC)

; transforming back to have EmaxRE with logit distribution (values between 0 and 1)
E0 = exp(tE0RE)/(1+exp(tE0RE))

; Maximal QoL (improved)
; transform the Emax fixed effect (EmaxFE) to tEmax (normally distribute)
tEmax = logit(EmaxFE)

; adding the random effects (RE) due to between-study variability and
tEmaxRE = tEmax + etaBSVEmax

; transforming back to have EmaxRE with logit distribution (values between 0 and 1)
Emax = exp(tEmaxRE)/(1+exp(tEmaxRE))


; Define QoL trajectory
SLP2 = SLP*0.001
Q = E0 - Emax * (1 - exp(-Kp * t)) + SLP2*t
; adding a saturation to avoid taking logit(0) (undefined) when t=0
Qsat = min(max(Q,0.01),0.99)
Qscore = Qsat*100

; transforming the effect Q in the same way as the data
pred = logit(Q)*sqrt(NOC)

OUTPUT:
output = {pred}
table ={Qscore, Emax, E0, SLP2}
```

### B. MBMA model Monolix settings.

```
<DATAFILE>

[FILEINFO]
file={path='../Combined_MBMA_QOL_Monolix_02JUN2025.csv'}
delimiter = comma
header={AID, STID, ID, PUBMEDID, NCT, TIMEWK, DV, DIS, DOSE, TRT, BSL, CFB, CFBVAR, CFBVARSTAT,
RESPONSE, RESPVAR, RESPVARSTAT, NOC, NTRT, ARM, AGE, MaleP, ECOG0P, ECOG1P, DSIP, DSIIP, DSIIIP,
DSIVP, TransDV, OID}

[CONTENT]
STID = {use=identifier}
TIMEWK = {use=time}
DIS = {use=covariate, type=categorical}
TRT = {use=covariate, type=categorical}
NOC = {use=regressor}
AGE = {use=covariate, type=continuous}
MaleP = {use=covariate, type=continuous}
ECOG0P = {use=covariate, type=continuous}
DSIVP = {use=covariate, type=continuous}
TransDV = {use=observation, type=continuous}
OID = {use=occasion}

<MODEL>

[COVARIATE]
input = {AGE, DSIVP, ECOG0P, MaleP, DIS, TRT}

DIS = {type=categorical, categories={'CRC', 'Endometrial', 'HNSCC', 'Melanoma', 'NSCLC', 'TNBC',
'Urothelial'}}
TRT = {type=categorical, categories={'Pembrolizumab', 'Placebo'}}

[INDIVIDUAL]
input = {E0FE_pop, EmaxFE_pop, Kp_pop, SLP_pop, etaBSVE0_pop, omega_etaBSVE0, etaBSVEmax_pop,
omega_etaBSVEmax, etaBTAVE0_pop, TRT, beta_EmaxFE_TRT_Placebo, gamma_SLP, beta_SLP_TRT_Placebo,
gamma_etaBTAVE0}

TRT = {type=categorical, categories={'Pembrolizumab', 'Placebo'}}

DEFINITION:
E0FE = {distribution=logitNormal, typical=E0FE_pop, no-variability}
EmaxFE = {distribution=logitNormal, typical=EmaxFE_pop, covariate=TRT, coefficient={0,
beta_EmaxFE_TRT_Placebo}, no-variability}
Kp = {distribution=logNormal, typical=Kp_pop, no-variability}
SLP = {distribution=logNormal, typical=SLP_pop, covariate=TRT, coefficient={0,
beta_SLP_TRT_Placebo}, varlevel=id*occ, sd=gamma_SLP}
etaBSVE0 = {distribution=normal, typical=etaBSVE0_pop, sd=omega_etaBSVE0}
etaBSVEmax = {distribution=normal, typical=etaBSVEmax_pop, sd=omega_etaBSVEmax}
etaBTAVE0 = {distribution=normal, typical=etaBTAVE0_pop, varlevel=id*occ, sd=gamma_etaBTAVE0}

[LONGITUDINAL]
input = {a}

file = 'test13.txt'

DEFINITION:
TransDV = {distribution=normal, prediction=pred, errorModel=constant(a)}

<FIT>
data = 'TransDV'
model = TransDV
```

```
<PARAMETER>
E0FE_pop = {value=0.65, method=MLE}
EmaxFE_pop = {value=0.05, method=MLE}
Kp_pop = {value=0.01, method=MLE}
SLP_pop = {value=0.7, method=MLE}
a = {value=1, method=MLE}
beta_EmaxFE_TRT_Placebo = {value=0, method=MLE}
beta_SLP_TRT_Placebo = {value=0, method=MLE}
etaBSVE0_pop = {value=0, method=FIXED}
etaBSVEmax_pop = {value=0, method=FIXED}
etaBTAVE0_pop = {value=0, method=FIXED}
gamma_SLP = {value=1, method=MLE}
gamma_etaBTAVE0 = {value=1, method=MLE}
omega_etaBSVE0 = {value=1, method=MLE}
omega_etaBSVEmax = {value=1, method=MLE}

<MONOLIX>

[TASKS]
populationParameters()
individualParameters(method = {conditionalMean, conditionalMode })
fim(method = Linearization)
logLikelihood(method = Linearization)

[PLOTS]
run = true
plots = {indfits = {selected = true}, parameterdistribution = {selected = true}, obspred = {selected
= true}, covariancemodeldiagnosis = {selected = true}, covariatemodeldiagnosis = {selected = true},
vpc = {selected = true}, residualsscatter = {selected = true}, residualsdistribution = {selected =
true}, randomeffects = {selected = true}, saemresults = {selected = true}}

[SETTINGS]
GLOBAL:
exportpath = 'test13'
```
